# Secular Trends in Incidence, Prevalence, and Survival of Pancreatic Cancer in the United Kingdom: A Population-Based Cohort Study from 2000 to 2021

**DOI:** 10.1101/2025.04.08.25325482

**Authors:** Maria Raffaella Romito, Eng Hooi Tan, Asieh Golozar, Talita Duarte-Salles, Antonella Delmestri, Wai Yi Man, Edward Burn, Daniel Prieto Alhambra, Carlos Guarner-Argente, Danielle Newby

## Abstract

**Background:** Pancreatic cancer is the seventh leading cause of cancer-related mortality worldwide and the fifth in the United Kingdom (UK), with incidence rates nearly matching mortality.

**Methods:** We conducted a population-based cohort study using Clinical Practice Research Datalink (CPRD) GOLD and Aurum databases including patients aged 18+ with at least one year of prior data, diagnosed between 2000 and 2021. We estimated incidence, period prevalence, and survival at one, five, and ten years, stratified by age, sex, and calendar year.

**Results:** A total of 10,116 (GOLD) and 20,127 (Aurum) patients were included (median age 73 years, 50% male). Crude incidence rose from 6.9 per 100,000 person-years in 2000 to 14.2 in 2021. Age-standardized rates were lower towards the end of the study. Period prevalence increased from 0.01% to 0.03% over the same period. Median survival was 0.38 (95% CI 0.36–0.39) years in GOLD and 0.39 (95% CI 0.38–0.40) in Aurum. One-, five-, and ten-year survival were 25.3–26.2%, 6.1–6.6%, and 3.8–4.0%, respectively, with no differences by sex. One-year survival improved from 22.5% (95% CI 20.2–25.1) in 2000–2004 to 28.4% (95% CI 26.5–30.5) in 2015–2019.

**Conclusions:** Pancreatic cancer incidence and prevalence are increasing in the UK. Rising incidence may partly reflect population aging. Survival remains poor, with only modest short-term improvements. Small improvements in survival highlight more research is needed to improve earlier diagnosis which will lead to better patient outcomes.

**KEY MESSAGES:**

- This study aimed to assess long-term trends in the incidence, prevalence, and survival of pancreatic cancer in the United Kingdom using two large primary care databases.
- We found that both incidence and prevalence of pancreatic cancer have increased between 2000 and 2021, while survival has remained poor, with only modest short-term improvements.
- These findings highlight the urgent need for strategies focused on early diagnosis and prevention, particularly considering population aging and the increasing burden of modifiable risk factors.

## INTRODUCTION

Pancreatic cancer is the seventh leading cause of cancer death worldwide in both sexes and the fifth in the United Kingdom (UK) ^1^, with a mortality rate (*n* = 466,000) almost equal to the incidence rate (*n* = 496,000) due to poor prognosis. Globally, stable or slight increasing incidence and mortality rates have been observed, especially in more developed countries ^2^, and it is projected to become the third most common cause of cancer-related mortality in the Europe, following lung and colorectal cancers ^3^

In the UK, pancreatic cancer incidence in both sexes have increased by 9% and mortality by 3%, with a ten-year survival rate of <1% that remained unchanged between 1970 and 2010 ^4^. A major reason for high mortality rates is late diagnosis, making most cases ineligible for curative surgery (up to 90%), resulting in a five-year net survival ^5^ rate of 4.0-7.9% from 2000 to 2014 in UK, lower than in other European countries ^6^. Poor survival can be attributed to non-specific symptoms in early disease that lead to delays in diagnosis. In fact, even symptomatic patients have poorer survival than asymptomatic individuals which are more likely to be diagnosed earlier, often incidentally ^7^. Unlike other cancers such as colorectal or breast, population screening program is not feasible except for some high-risk groups, due to its lower prevalence and lack of validated early tests ^8^.

Conversely, incidence and mortality could be reduced by addressing modifiable risk factors, including smoking, alcohol, obesity, diabetes, and other diet, which have become more prevalent in recent decades ^9^.

There is a lack of up-to-date comprehensive assessment of pancreatic cancer trends in the UK, and with changes in risk factors in recent decades, continuous epidemiological evaluation is necessary. Analysis of pancreatic cancer epidemiology may be the key to help further understand its aetiology and thus support effective prevention strategy. This study aims to describe pancreatic cancer burden and trends in incidence, prevalence, and survival from 2000 to 2021 using two UK primary care databases.

## METHODS

### Study design, setting, and data sources

This population cohort study used routinely collected primary care data from the UK. People with a diagnosis of pancreatic cancer and a denominator cohort were identified from Clinical Practice Research Datalink (CPRD) GOLD to estimate overall survival, incidence, and prevalence. We repeated the analysis using CPRD Aurum to compare the results with GOLD. Both databases are broadly representative of the UK population and contain pseudonymized patient-level information on demographics, lifestyle, clinical diagnoses, prescription, and preventative care^10^. Both databases were mapped to the Observational Medical Outcomes Partnership (OMOP) Common Data Model (CDM) ^11^ ^12^. The use of CPRD data was approved by the Research Data Governance (RDG)(Protocol 22_001843) using data cuts 2022/07 and 2021/06 for GOLD and Aurum respectively.

### Study participants

Participants were 18 years or older with at least one year of prior history in the database. For the incidence and prevalence analysis, the study cohort consisted of individuals present in the database from 1st January 2000. These individuals were followed up to whichever came first: the cancer outcome of interest, exit from the database, date of death, or the or the end of study (December 31, 2021, for GOLD, and, due to data availability of latest extraction, December 31, 2019, for Aurum). For the survival analysis, individuals were followed up from the date of their cancer diagnosis to either date of death, exit from database or end of the study period. Any patients whose death and cancer diagnosis occurred on the same date were excluded.

### Pancreatic cancer definition

We used Systematized Nomenclature of Medicine Clinical Terms (SNOMED CT) diagnostic codes to identify pancreatic cancer events. Diagnostic codes indicative of non-malignant or metastatic disease were excluded (apart from prevalence analysis). The pancreatic cancer definition was reviewed by clinicians with oncology, primary care, and real-world data expertise (supplementary table S1). For survival analysis, mortality was defined as all-cause mortality based on records of date of death. OMOP-based computable phenotypes are available, together with all analytical code at on Github to enable reproducibility (https://github.com/oxford-pharmacoepi/EHDENCancerIncidencePrevalence).

### Statistical methods

The characteristics of patients with pancreatic cancer were summarised, with median (IQR) used for continuous variables and counts and percentages used for categorical variables.

Crude overall and annual incidence rates (IR) for pancreatic cancer were calculated from 2000 to 2021. For incidence, the number of events, the observed time at risk, and the incidence rate per 100,000 person years were summarised along with 95% confidence intervals (95% CI).

Age-standardized IRs were calculated using the 2013 European Standard Population (ESP2013) ^13^. Age-standardized rates for GOLD were compared to national cancer statistics^14^.

Period prevalence was calculated on 1^st^ January for the years 2000 to 2021, with the number of pancreatic cancer patients as the numerator. The denominator included patients on 1^st^ January in the respective years for each database. The number of events, and prevalence (%) were summarised along with 95% CI.

For survival analysis, we used the Kaplan-Meier (KM) method to estimate the overall survival with 95% confidence intervals. We estimated median survival and survival one, five, and ten years after diagnosis. All results were stratified by age and sex. Additionally, for CPRD GOLD, we additionally stratified by calendar time of cancer diagnosis (2000-2004, 2005-2009, 2010-2014, 2015-2019 and 2020-2021). To avoid re-identification, we do not report results with fewer than five cases.

For Aurum, the same statistical analyses, except for stratification by calendar time of cancer diagnosis, which was conducted in GOLD only.

The statistical software R version 4.2.3 was used for analyses. For calculating incidence and prevalence, we used the IncidencePrevalence R package ^15^. For survival analysis we used the survival R package ^16^.

## RESULTS

### Patient Populations and characteristics

Overall, there were 11,388,117 eligible patients, with at least one year of prior history identified from January 2000 to December 2021 from GOLD. Attrition tables can be found in supplement S2. A summary of baseline patient characteristics of those with a diagnosis of pancreatic cancer is shown in Table 1.

**Table 1:** Baseline characteristics of pancreatic cancer patients at the time of diagnosis for CPRD GOLD.

| **Database** | **CPRD GOLD** |
| --- | --- |
| **Number of patients** | 10116 |
| **Sex: Male (N[%])** | 5035 (49.8%) |
| **Age (Median [IQR])** | 73 (64 to 80) |
| **Age Groups N (%)** |  |
| 18-29 | 6 (0.1%) |
| 30-39 | 35 (0.3%) |
| 40-49 | 314 (3.1%) |
| 50-59 | 1136 (11.2%) |
| 60-69 | 2546 (25.2%) |
| 70-79 | 3316 (32.8%) |
| 80-89 | 2337 (23.1%) |
| 90+ | 426 (4.2%) |
| **Prior history, days** |  |
| median [IQR] | 3776 (2,103 to 5,489) |
| **Smoking Status (any time 5 years prior)** |  |
| Nonsmoker | 4326 (42.8%) |
| Former Smoker | 86 (0.9%) |
| Current Smoker | 2296 (22.7%) |
| Missing | 3408 (33.7%) |
| **General conditions (any time prior)** |  |
| Atrial fibrillation | 645 (6.4%) |
| Cerebrovascular disease | 649 (6.4%) |
| Chronic obstructive lung disease | 743 (7.3%) |
| Coronary arteriosclerosis | 110 (1.1%) |
| Dementia | 167 (1.7%) |
| Depressive disorder | 1229 (12.1%) |
| Diabetes mellitus | 2324 (23.0%) |
| Gastroesophageal reflux disease | 340 (3.4%) |
| Gastrointestinal haemorrhage | 775 (7.7%) |
| Heart failure | 321 (3.2%) |
| Hyperlipidemia | 1005 (9.9%) |
| Hypertensive disorder | 2867 (28.3%) |
| Ischemic heart disease | 1106 (10.9%) |
| Osteoarthritis | 2346 (23.2%) |
| Peripheral vascular disease | 238 (2.4%) |
| Pulmonary embolism | 174 (1.7%) |
| Renal impairment | 1399 (13.8%) |
| Venous thrombosis | 670 (6.6%) |
| Type II diabetes | 2065 (20.4%) |

IQR: interquartile range

Pancreatic cancer shows a balanced sex distribution, with a median age of 73 years old across both databases. The highest percentage of patients were those aged 70-79 years old (33% of diagnosed patients). Baseline characteristics for Aurum were similar to GOLD (Supplement S3).

### Overall and annualised crude incidence rates with age and sex stratifications

The overall IR of pancreatic cancer from 2000 to 2021 was 11.5 (11.3 to 11.8) per 100000 person-years for GOLD. Males and females had similar overall IR values (11-12 per 100000 person-years) for both databases. Annualised IRs increased across the study period (Figure 1). Age standardization of IR showed the same trends but with a smaller increase over time with slight attenuation towards the end of study period (Supplement S4). Comparison with national cancer registry data from England showed our estimates were slightly lower with same underlying trends (Supplement S5). All study results for this study can be found in a user-friendly interactive web application: https://dpa-pde-oxford.shinyapps.io/IncidencePrevalenceCancers/.

**Figure 1:**
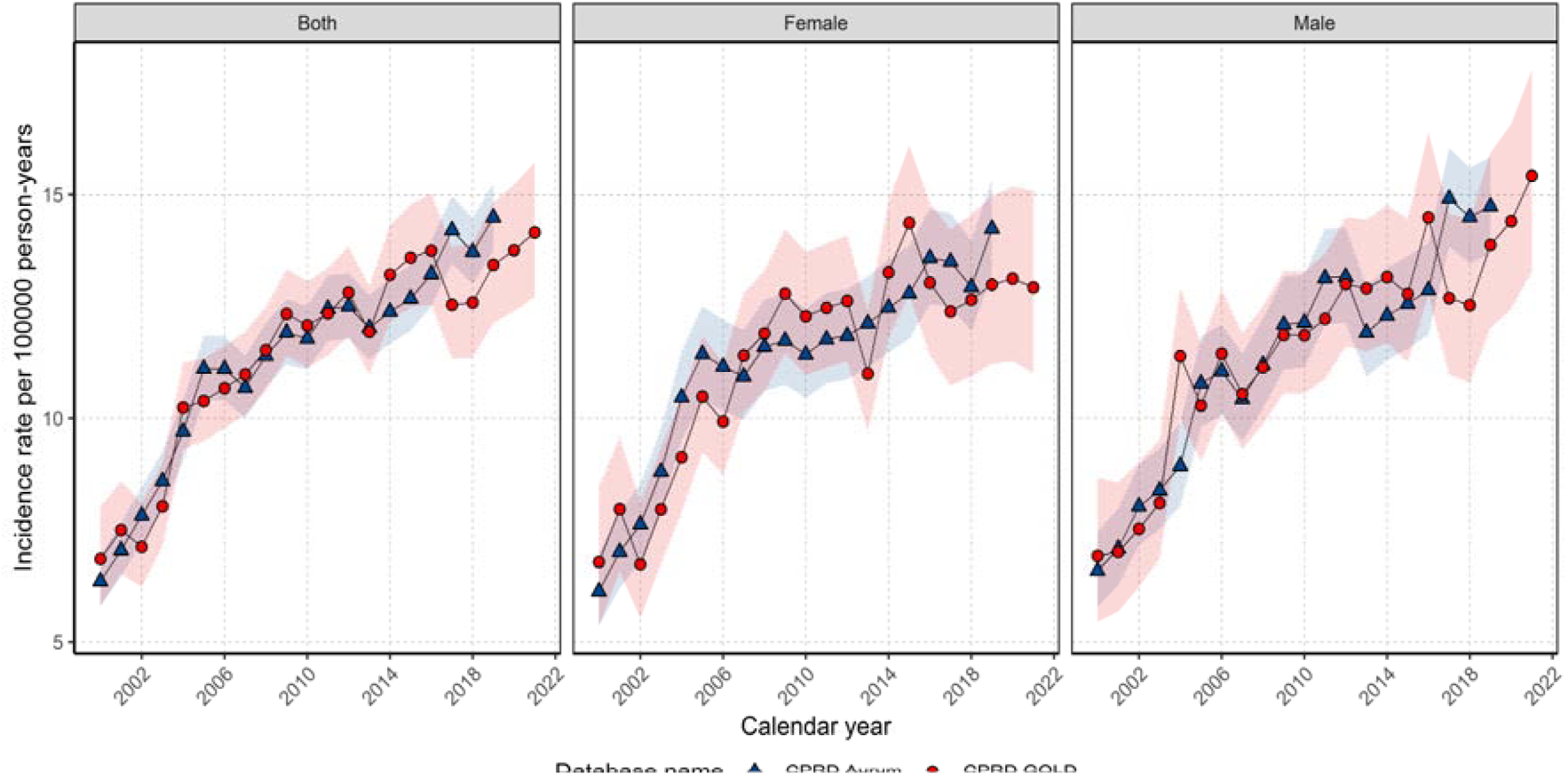
Crude incidence rates for pancreatic cancer stratified by database and sex. Overall crude IRs were higher with increasing age up to 80–89 years. Those aged 18 to 29 had overall IRs of 0.04 per 100000 person years and those aged 80–89-years with IRs of 38.2-44.1 in Aurum and GOLD respectively (Supplement S6) with similar trends for both sexes.

Annualised IRs for pancreatic cancer for each age group are shown in Figure 2. There were not enough data points to assess annualised trends for those ages 18 to 39 years of age. All age groups apart from those aged 60-69 years of age show an increase in IRs across the study period. For those aged 60-69 years of age, annualised IRs increased from 2000 to 2010 and then remained stable to the end of the study period. Stratification on both sex and age group showed similar trends in Figure 2 with minimal sex differences between annualised IRs for most age groups apart from those aged 60-79 years of age where there were slightly higher IRs for males (Supplement S7).

**Figure 2:**
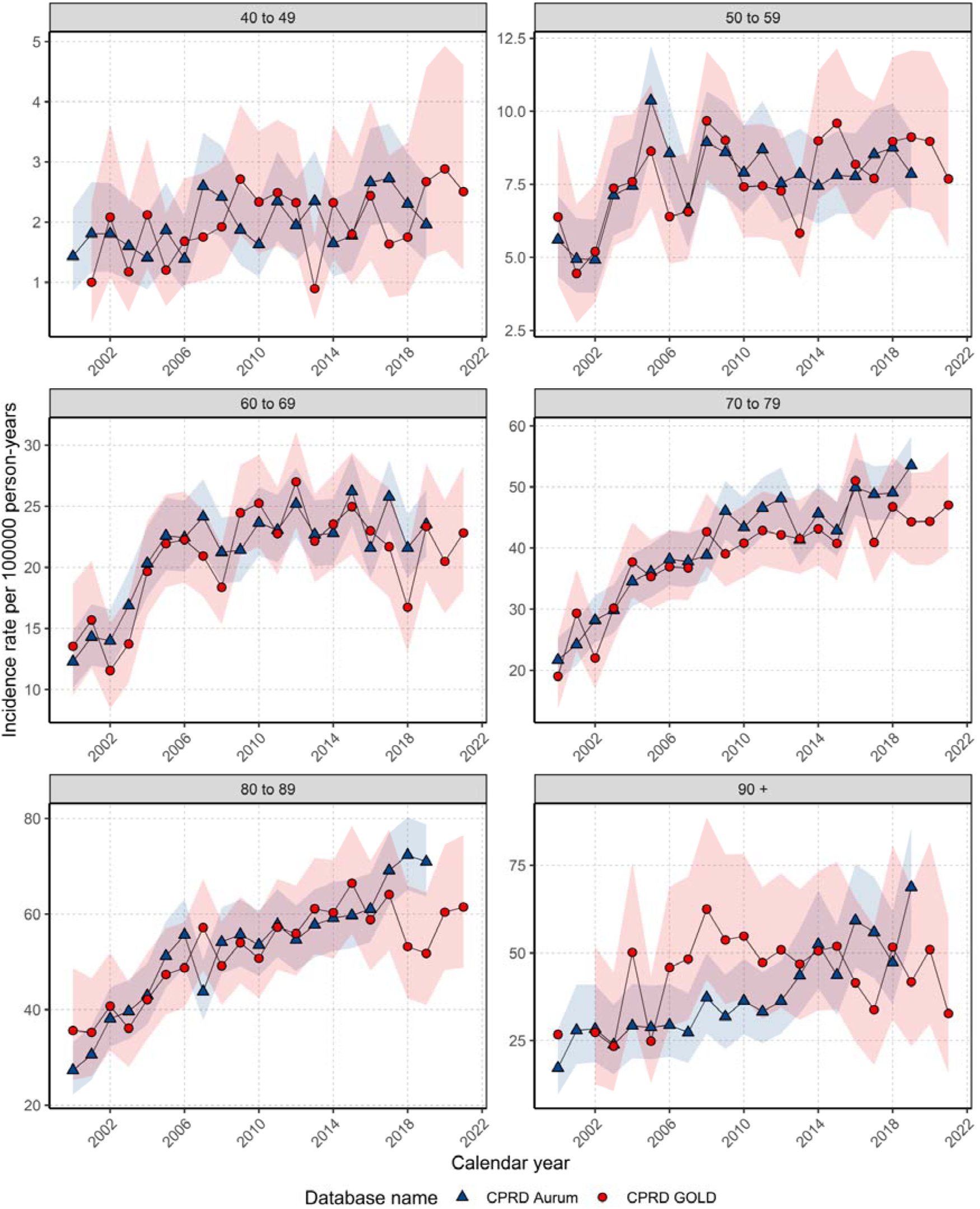
Incidence rates for pancreatic cancer stratified by database and age group. Period prevalence with database, age, and sex stratifications. Crude PP for pancreatic cancer in 2021 was 0.028% (0.026% to 0.030%) for GOLD. PP in 2019 was similar across both databases (0.026-0.028%) with minimal sex differences. PP increased over the study period (Figure 3). In GOLD, PP increased 2.8-fold from 2000 to 2021 with no sex differences with similar results in Aurum.

**Figure 3:**
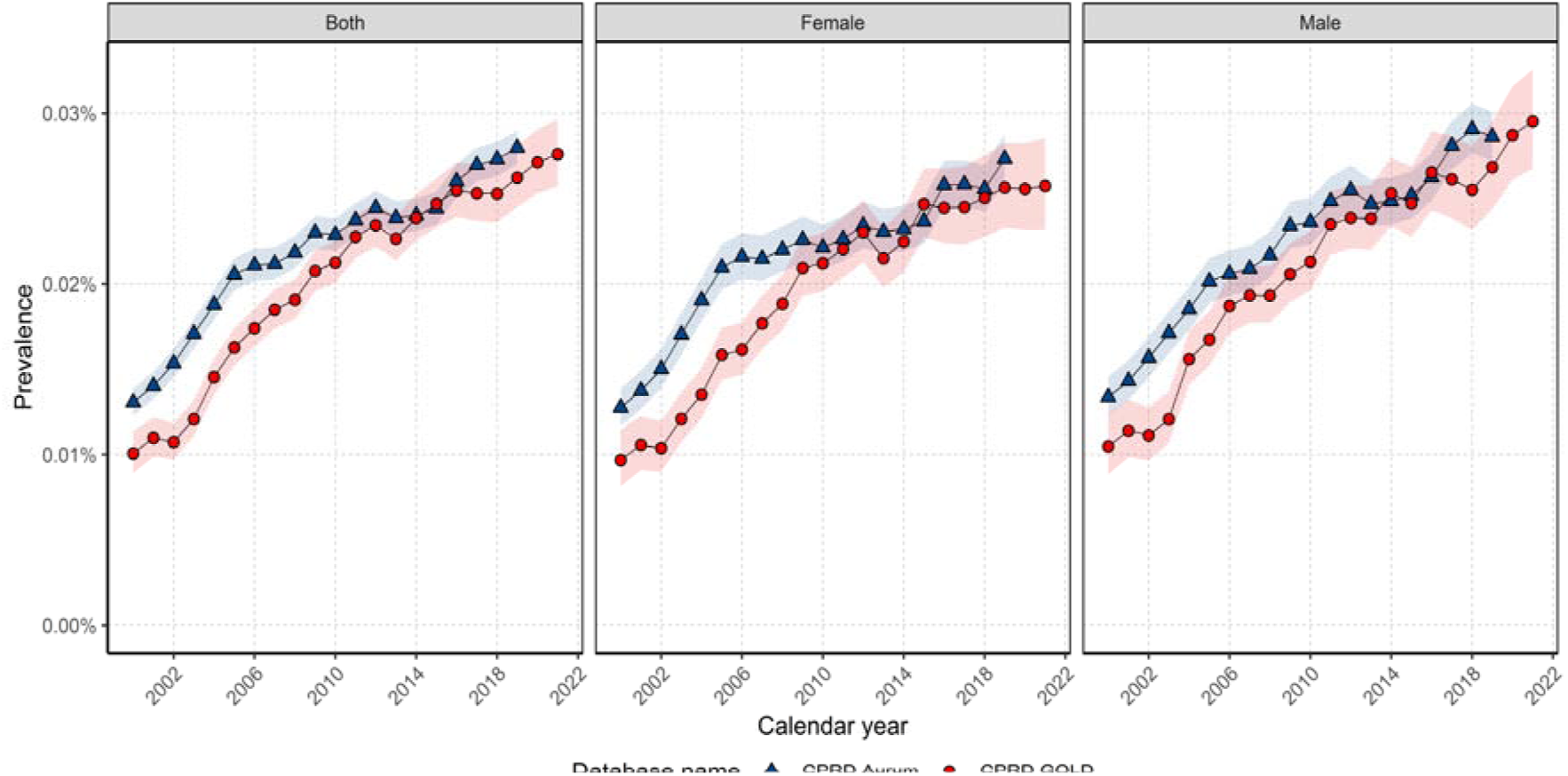
Crude period prevalence for stratified by database and sex. PP in 2019 was highest in those aged 80-89 years of age (0.13% Aurum, 0.11% GOLD). Trends in annualised PP show increases in PP across the study period for all age groups apart from those aged 60-69 years of age, with PP starting to decrease from 2016 (Figure 4). Stratification on both sex and age group showed similar trends in Figure 4. For those aged 50-79, annualised PP were slightly higher in males and for all other age groups there were no sex differences (Supplement S8).

**Figure 4:**
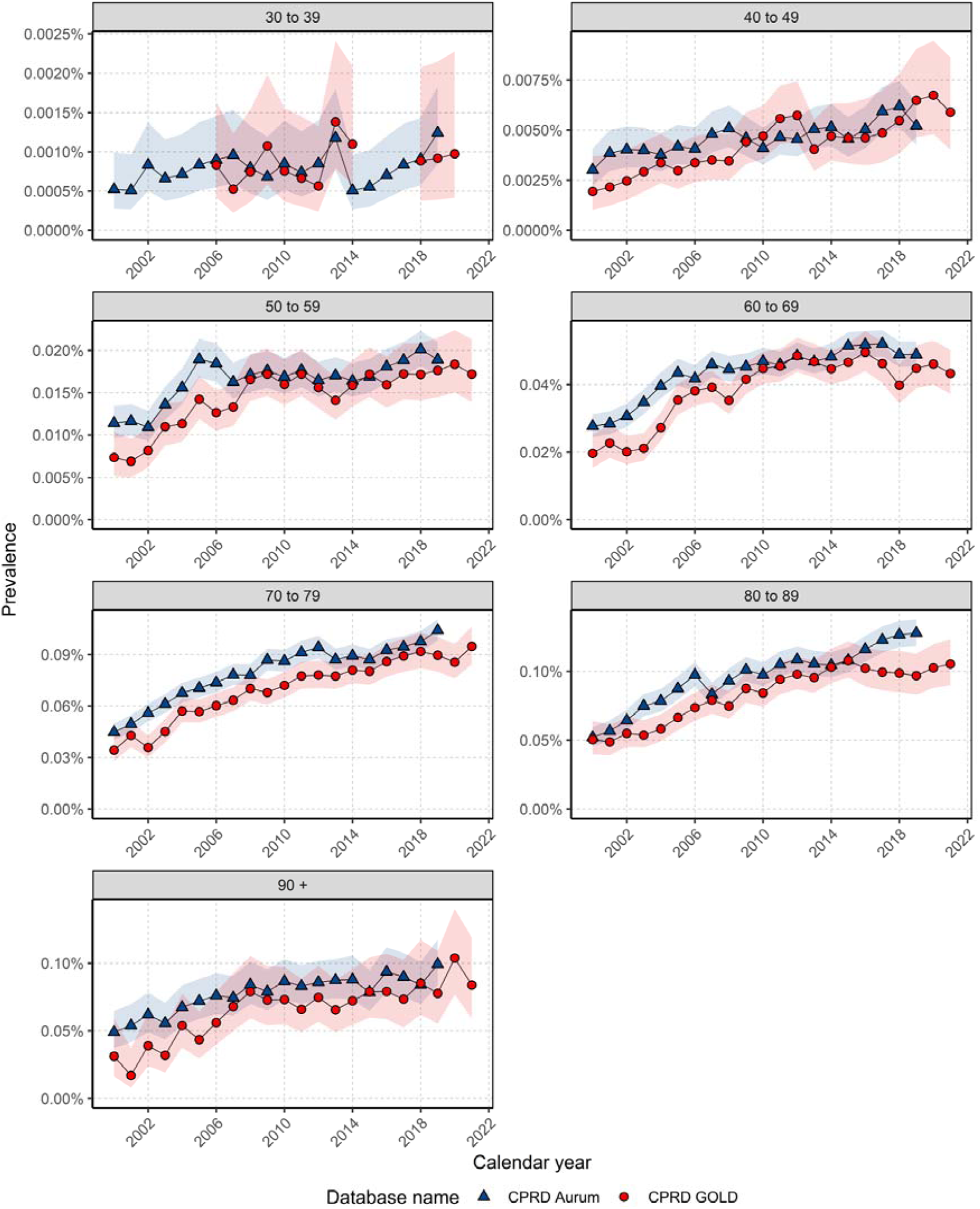
Annualised period prevalence for pancreatic cancer stratified by database and age group.

### Overall survival with age, sex, and calendar year stratification

There were 9,770 and 19,629 patients with 8,430 and 16,806 deaths with a median follow-up of 0.36 (IQR 0.36 - 0.39) and 0.34 years (0.12 – 0.89) years for GOLD and Aurum respectively. (Supplement S9). Median survival for the whole population was 0.38 (0.36 - 0.39) years and 0.39 (95% CI 0.38 - 0.40) years. Survival after one, five and ten years after diagnosis was 25.3%, 6.1% and 3.8% for GOLD with similar values for Aurum. Males and females had similar median survival of around 0.38 years across both databases with short- and long-term survival showing no sex differences (Supplement S10). When stratifying by age group, median, short- and long-term survival decreased with age from 30 years of age for both databases with the same result when stratifying by sex (Table 2).

**Table 2:** Survival rates of pancreatic cancer stratified by database and age group.

| **Age Group** | **One year survival (%)** |  | **Five-year survival (%)** |  | **Ten-year survival (%)** |  |
| --- | --- | --- | --- | --- | --- | --- |
|  | Aurum | GOLD | Aurum | GOLD | Aurum | GOLD |
| 18-29 | 75.5 (54.9 -100.0) | - | - | - | - | - |
| 30-39 | 62.6 (52.8 - 74.3) | 51.4 (37.3 – 71.0) | 40.5 (29.8 - 55.1) | 35.5 (22.4 - 56.3) | 27.8 (17.0 - 45.4) | - |
| 40-49 | 41.4 (37.6 - 45.5) | 41.8 (36.5 - 48.0) | 18.3 (15.2 -22.0) | 17.0 (12.9 - 22.5) | 10.3 (7.6 - 14.0) | 9.9 (6.1 - 16.1) |
| 50-59 | 36.2 (34.2 - 38.4) | 35.3 (32.5 - 38.3) | 10.2 (8.8 - 11.8) | 10.7 (8.8 - 13.0) | 6.7 (5.4 - 8.2) | 5.5 (3.9 - 7.8) |
| 60-69 | 31.2 (29.8 - 32.6) | 28.6 (26.8 - 30.5) | 7.6 (6.8 - 8.6) | 6.8 (5.7 - 8.1) | 4.9 (4.13 - 5.8) | 4.3 (3.4 - 5.6) |
| 70-79 | 25.2 (24.1 - 26.3) | 24.5 (23.0 - 26.1) | 5.5 (4.9 - 6.2) | 4.8 (3.9 - 5.8) | 3.7 (3.1 - 4.4) | 3.4 (2.7 - 4.4) |
| 80-89 | 16.5 (15.4 - 17.7) | 16.8 (15.3 - 18.6) | 3.1 (2.5 - 3.8) | 3.3 (2.5 - 4.4) | 1.0 (0.6 - 1.8) | 1.8 (1.1 - 3.0) |
| 90+ | 11.2 (9.0 - 13.8) | 11.2 (8.3 - 15.2) | 3.0 (1.8 - 5.0) | 1.7 (0.6 - 5.0) | 0.6 (0.2 - 2.4) | - |

Results for 18–29-year-olds not reported due to small sample numbers (< 5 cases)

For GOLD, median survival did not change over calendar time however, for short-term survival there was a small improvement in survival (Figure 5). For those diagnosed in 2000-2004 survival was 22.5% (20.2 - 25.1) whereas those diagnosed in 2015-2019 survival was 28.4 (26.5 - 30.5). For different age groups there was no clear pattern over calendar time for short- and long-term survival (Supplement S11).

**Figure 5:**
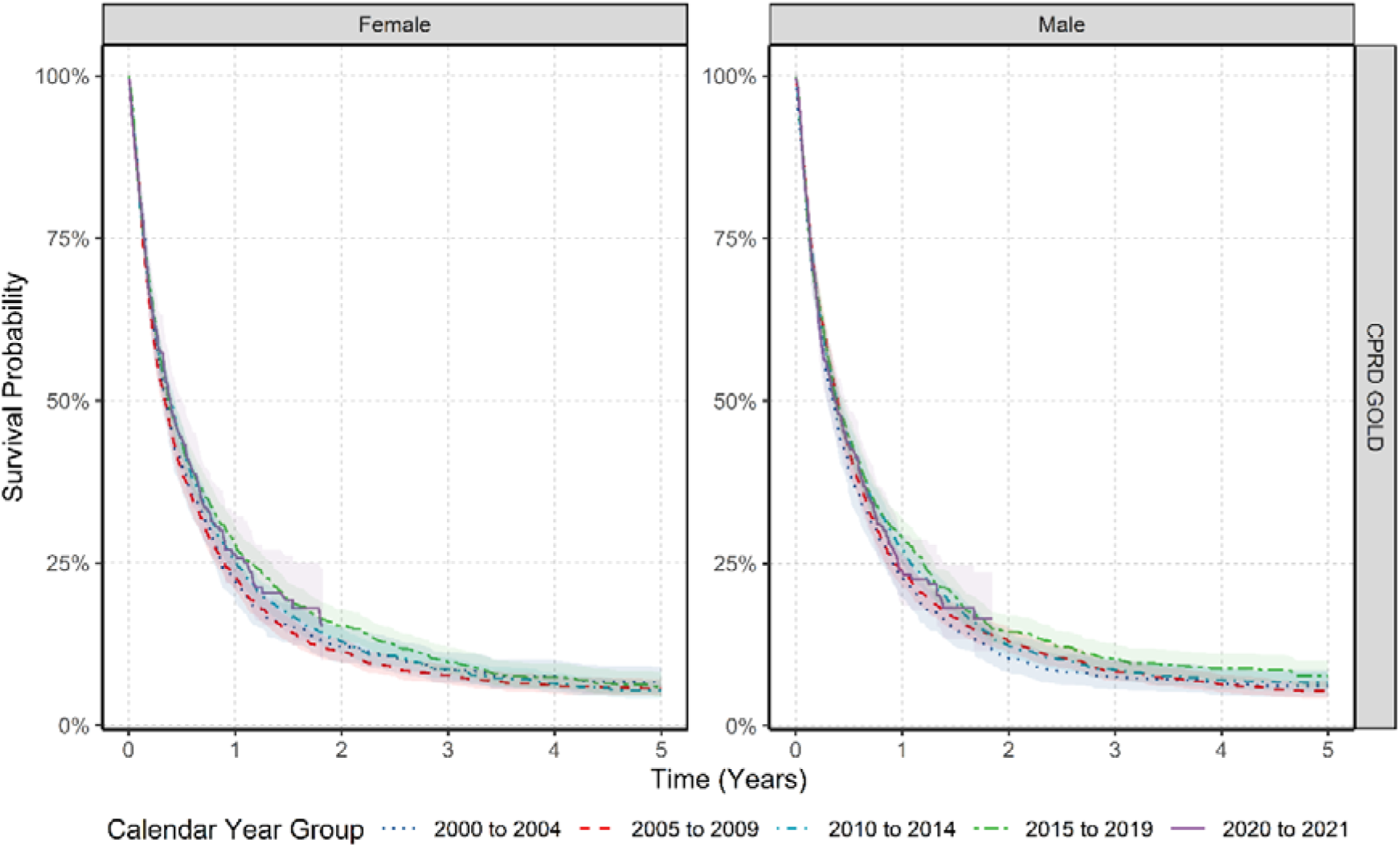
Kaplan-Meier survival curve of pancreatic cancer stratified by sex, and calendar year of diagnosis (2000-2004, 2005-2009, 2010-2014, 2015-2019 and 2020-2021) for CPRD GOLD.

## DISCUSSION

This paper provides a comprehensive study of trends in pancreatic cancer incidence, prevalence, and survival in the UK. Overall, the findings still paint a troubling picture: from 2000 to 2021 incidence and prevalence increased in both sexes, with a slight attenuation due to ageing populations over time particularly for females. Median survival remained just 0.38 years, with only a small improvement in one year survival over the study period.

Incidence rates reported here are slightly lower but follow similar trends to those from the National Cancer Registration and Analysis Service in England ^14^. The discrepancy likely reflects pancreatic cancer being more commonly diagnosed in hospital than in primary care. Comparing crude IR with age-standardized rates reveals less pronounced trends over time, particularly among females, suggesting that aging population structure, alongside risk factors, may partly explain the rise in cases.

We observed minimal sex differences in incidence, prevalence and survival, contradicting previous studies showing higher rate in males. However, upon age standardization for incidence, sex differences in line with previous studies was observed indicating population age structures of the disease have changed over time ^17^. Higher male rates reflect greater prevalence of risk factors, like tobacco, alcohol, visceral obesity, occupational and environmental factors, as well as a genetic susceptibility ^17^. Whereas for females, protective hormonal effects could also explain lower incidence and prevalence ^18^

Regarding survival, studies show conflicting results. A population-based study using national cancer registry data showed that females aged 56-74 with metastatic pancreatic cancer had a marginally longer median survival ^18^. However, the observed survival differences may lack clinically relevance. Conversely, another study showed females with stage I-III pancreatic cancer had poorer median survival however, their older age could explain the difference ^19^. This study also found that females were less likely to receive curative treatments, despite similar tumour characteristics, suggesting potential gender biases in treatment allocation, which may contribute to poorer survival ^19^. Additionally, a recent European study including electronic health records and cancer registries showed minimal differences between sexes for all survival outcomes ^20^ indicating sex differences maybe specific for certain subgroups and treatments ^21^.

Pancreatic cancer mainly affects older adults, with incidence peaking from age 70 ^17^. Additionally, we observed a steady rise in incidence in those over 40 years of age, likely due to longer exposure to risk factors compared to younger age groups. These, combined with genetic predisposition, may lead to mutation accumulation driving cancer development ^22^ Evidence from the UK Biobank study highlighted that adherence to a healthy lifestyle reduce pancreatic cancer risk regardless of age, with the lowest risk in individual under 60, indicating the importance of adopting healthy behaviours earlier in life, regardless of genetic risk ^23^. Furthermore, our age standardized results show similar trends with crude incidence rates with only slight attenuation in later years particularly for females indicating factors other than age, could be contributing to the increases in cancer cases over the study period more for males than females. The adoption of a healthy lifestyle in recent years could also explain our stable incidence rates from 2010 in those aged 50-59 and 60-69 years, resulting in a subsequent reduction in prevalence. Societal policies promoting healthy habits, such as exercising until a more advanced age ^24^ and smoking cessation ^25^, may modestly delay disease onset. Older age at diagnoses also impacts survival outcomes due to limited therapeutic options in elderly patients who, due to comorbidities, may not be candidates for invasive treatments^26^. However, some studies suggest that eligible older patients may receive less treatment, possibly due to access inequities or decision-making differences ^27^

Diabetes mellitus (DM) play an important role in the risk and manifestation of pancreatic cancer. It is therefore unsurprising we observed a large proportion of patients with pre-existing DM particularly type 2. Evidence suggests that not only having type 2 DM for more than five years increase pancreatic cancer risk ^7^ but those over 50 with new-onset diabetes have up to eight times the risk of pancreatic ductal adenocarcinoma ^28^ indicating type 2 DM can be both a paraneoplastic manifestation and a risk factor^29^. With the worryingly increases in obesity and type 2 DM due to poor diet and sedentary lifestyles in recent decades, implementing pancreatic cancer screening for all diabetic patients is not cost-effective, given the high prevalence of DM and low pancreatic cancer prevalence. However, UK, primary healthcare guidelines, issued by the National Institute for Health and Care Excellence (NICE), now recommend urgent abdominal imaging for individuals over 60 with newly diagnosed type II diabetes and unexplained weight loss ^30^.

Pancreatic cancer survival is low in line with both national ^31^ ^32^ and international studies ^33^ ^34^ which have also shown that pancreatic cancer mortality has remained stable or with slight improvements in the short term. Poor survival rates are largely due to the late diagnosis of the disease, as symptoms like abdominal or back pain, diarrhoea, constipation, dyspepsia, nausea, vomiting and newly developed diabetes are nonspecific and individually carry low pancreatic cancer risk, unless significant weight loss is present ^35^. Conversely, jaundice has the highest associated risk, especially in individuals over 40, as highlighted in NICE guidelines ^30^.

Additionally, late diagnosis results in limited curative treatment options with only 10% of patients eligible for surgical resection ^36^. However, some improvements have been seen with neoadjuvant therapy for resectable and locally advanced pancreatic cancer which could explain modest short term survival improvements. Limited results have been seen with immunological control therapies, despite their success in treating other solid cancers ^37^. Low funding and support, hinders progress in developing effective treatments for advanced stages of the disease ^38^.

The main strength of this study is the use of two large primary care databases with good coverage across the whole of the UK. Another strength of our study is the inclusion of a complete study population database for estimating incidence and prevalence as well as the ability to characterise comorbidities and medication use before diagnosis. In contrast, cancer registry studies extrapolate using a denominator from national population statistics, potentially introducing biases. Finally, the high validity and completeness of mortality data, with over 98% accuracy when compared to national mortality records ^39^ allowed us to understand the impact of calendar time on overall survival, one of the key outcomes in cancer care. However, our study had limitations. we used primary care data without linkage to the cancer registry, potentially leading to misclassification and delayed recording of diagnoses. However, previous validation studies have shown high accuracy and completeness of cancer diagnoses in primary care records ^40^ Furthermore, our use of primary care records precluded us from studying tumour histology, genetic mutations, staging or cancer therapies which can all impact survival.

## CONCLUSION

The rising incidence of pancreatic cancer is likely associated with an aging population and the increase in risk factors, particularly among males, emphasizing the need to identify individuals at risk and implement effective risk reduction strategies for future generations. Unlike other cancers, pancreatic cancer has seen no significant improvement in long-term survival rates over the past 20 years, with only a modest increase in short-term survival. Achieving future improvements in survival rates will require greater investment in public education, screening population models, advancements in diagnostic techniques, and enhanced therapeutic strategies.

## Data Availability

This study is based in part on data from the Clinical Practice Research Datalink (CPRD) obtained under licence from the UK Medicines and Healthcare products Regulatory Agency. The data is provided by patients and collected by the NHS as part of their care and support. The interpretation and conclusions contained in this study are those of the author/s alone. Patient level data used in this study was obtained through an approved application to the CPRD (application number 22_001843) and is only available following an approval process to safeguard the confidentiality of patient data. Details on how to apply for data access can be found at https://cprd.com/data-access.

## CONTRIBUTIONS

All authors were involved in the study conception and design, interpretation of the results, and the preparation of the manuscript. DN carried out data analysis for the manuscript. AG reviewed the clinical code list used in this study. RR and DN wrote the initial draft of the manuscript with EB, CGA, DPA. AD and WYM implemented the data curation, data harmonisation, data quality tests and assessment. DN, EB, AD, WYM and DPA had access to the CPRD data. All authors were involved in the interpretation of the results, critically reviewed the final manuscript, and gave consent for publication.

## ETHICAL APPROVAL

Patient level data was obtained by CPRD’s Research Data Governance Process (application number 22_001843). Individual patient consent was not required, as CPRD data are de-identified and provided under ethical approval from the UK Health Research Authority (HRA) and the NHS Health and Social Care Research Ethics Committee. The study complies with all relevant ethical regulations, including the principles outlined in the Declaration of Helsinki.

## ABBREVIATIONS

IR: incidence rate
PP: period prevalence
UK: United Kingdom
DM: diabetes mellitus

