## Supplement for "Secular Trends in Incidence, Prevalence, and Survival of Pancreatic Cancer in the United Kingdom: A Population-Based Cohort Study from 2000 to 2021"

### **S1: Clinical codelists for pancreatic cancer**

The clinical codelists used for pancreatic cancer is listed in the table below with the corresponding SNOMED concept ID, OMOP concept ID and concept description. Only diagnosis records alone were used to identify cancer outcome for this study. Different codelists were created for incident and prevalent definitions of pancreatic cancer. We developed concept definitions using ATLAS, the OHDSI open-source platform (<https://github.com/OHDSI/atlas>). Clinical adjudicators reviewed the cohort definitions and associated concept sets.

| **Concept Id** | **Concept SNOMED Code** | **Concept Description** | **Code used for** |
| --- | --- | --- | --- |
| 4180793 | 363418001 | Malignant tumor of pancreas | Incidence and prevalence |
| 4178967 | 363419009 | Malignant tumor of head of pancreas | Incidence and prevalence |
| 4094866 | 187793004 | Malignant tumor of pancreatic duct | Incidence and prevalence |
| 4095436 | 187792009 | Malignant tumor of tail of pancreas | Incidence and prevalence |
| 4092072 | 187791002 | Malignant tumor of body of pancreas | Incidence and prevalence |
| 4095437 | 187794005 | Malignant tumor of Islets of Langerhans | Incidence and prevalence |
| 25486 | 93843007 | Primary malignant neoplasm of islets of Langerhans | Incidence and prevalence |
| 199754 | 372003004 | Primary malignant neoplasm of pancreas | Incidence and prevalence |
| 432843 | 94082003 | Primary malignant neoplasm of tail of pancreas | Incidence and prevalence |
| 433423 | 93939009 | Primary malignant neoplasm of pancreatic duct | Incidence and prevalence |
| 434293 | 93715005 | Primary malignant neoplasm of body of pancreas | Incidence and prevalence |
| 440649 | 372119009 | Primary malignant neoplasm of head of pancreas | Incidence and prevalence |
| 4110585 | 254612002 | Carcinoma of endocrine pancreas | Incidence and prevalence |
| 4111024 | 255088001 | Malignant tumor of exocrine pancreas | Incidence and prevalence |
| 4112734 | 254611009 | Malignant tumor of endocrine pancreas | Incidence and prevalence |
| 4157459 | 372142002 | Carcinoma of pancreas | Incidence and prevalence |
| 4178960 | 363369002 | Carcinoma of tail of pancreas | Incidence and prevalence |
| 4181331 | 363368005 | Carcinoma of body of pancreas | Incidence and prevalence |
| 4209933 | 326072005 | Carcinoma of head of pancreas | Incidence and prevalence |
| 4340498 | 235966007 | Cystadenocarcinoma of pancreas | Incidence and prevalence |
| 36683250 | 780821007 | Invasive intraductal papillary-mucinous carcinoma of pancreas | Incidence and prevalence |
| 36713362 | 681621000119105 | Primary adenocarcinoma of body of pancreas | Incidence and prevalence |
| 36713363 | 681721000119103 | Primary adenocarcinoma of head of pancreas | Incidence and prevalence |
| 37204187 | 782697005 | Solid pseudopapillary carcinoma of pancreas | Incidence and prevalence |
| 37204852 | 783771003 | Acinar cell carcinoma of pancreas | Incidence and prevalence |
| 37206235 | 785879009 | Mucinous cystadenocarcinoma of pancreas | Incidence and prevalence |
| 37311469 | 792907004 | Pancreatic ductal adenocarcinoma | Incidence and prevalence |
| 37395837 | 715414009 | Familial malignant neoplasm of pancreas | Incidence and prevalence |
| 42872399 | 1651000119109 | Primary adenocarcinoma of pancreas | Incidence and prevalence |
| 45763891 | 700423003 | Adenocarcinoma of pancreas | Incidence and prevalence |
| 4201015 | 314999005 | Metastasis from malignant tumor of pancreas | Prevalence only |
| 4201481 | 314964006 | Local recurrence of malignant tumor of pancreas | Prevalence only |

### **S2: Population attrition showing eligible patients for study from each database.**

| **N** | **Reason** | **N excluded** | **Database** |
| --- | --- | --- | --- |
| 39999011 | Starting population |  | Aurum |
| 39999011 | Missing year of birth | 0 |  |
| 39999011 | Missing sex | 0 |  |
| 34833388 | Cannot satisfy age criteria during the study period based on year of birth | 5165623 |  |
| 29190480 | No observation time available during study period | 5642908 |  |
| 29190480 | Doesn't satisfy age criteria during the study period | 0 |  |
| 25483313 | Prior history requirement not fulfilled during study period | 3707167 |  |
| 24340860 | No observation time available after applying age and prior history criteria | 1142453 |  |
| 24340860 | Starting analysis population |  |  |
| 24340860 | Estimating prevalence |  |  |
| 24339673 | Excluded due to prior event (do not pass outcome washout during study period) | 1187 |  |
| 24339673 | Estimating incidence |  |  |
| 20127 | With a cancer diagnosis |  |  |
| 19629 | Cancer diagnosis not on same date as death | 498 |  |
| 19629 | Estimating survival |  |  |
| 17054819 | Starting population |  | GOLD |
| 17054819 | Missing year of birth | 0 |  |
| 17054819 | Missing sex | 0 |  |
| 15210165 | Cannot satisfy age criteria during the study period based on year of birth | 1844654 |  |
| 13978229 | No observation time available during study period | 1231936 |  |
| 13978229 | Doesn't satisfy age criteria during the study period | 0 |  |
| 12254874 | Prior history requirement not fulfilled during study period | 1723355 |  |
| 11388117 | No observation time available after applying age and prior history criteria | 866757 |  |
| 11388117 | Starting analysis population |  |  |
| 11388117 | Estimating prevalence |  |  |
| 11387716 | Excluded due to prior event (do not pass outcome washout during study period) | 401 |  |
| 11387716 | Estimating incidence |  |  |
| 10116 | With a cancer diagnosis |  |  |
| 10116 | Cancer diagnosis not on same date as death | 346 |  |
| 9770 | Estimating survival |  |  |

### **S3: Baseline characteristics of pancreatic cancer patients at the time of diagnosis for CPRD Aurum.**

| **Database** | **CPRD Aurum** |
| --- | --- |
| **Number of patients** | 20127 |
| **Sex: Male (N[%])** | 10123 (50.30%) |
| **Age (Median [IQR])** | 73 (64 to 80) |
| **Age Groups N (%)** | |
| 18-29 | 14 (0.10%) |
| 30-39 | 93 (0.50%) |
| 40-49 | 656 (3.30%) |
| 50-59 | 2214 (11.00%) |
| 60-69 | 4873 (24.20%) |
| 70-79 | 6661 (33.10%) |
| 80-89 | 4748 (23.60%) |
| 90+ | 868 (4.30%) |
| **Prior history, days** | |
| median [IQR] | 6828 (3,537 to 11,221) |
| **General conditions (any time prior)** | |
| Atrial fibrillation | 1489 (7.4%) |
| Cerebrovascular disease | 1439 (7.1%) |
| Chronic liver disease | 101 (0.5%) |
| Chronic obstructive lung disease | 1527 (7.6%) |
| Coronary arteriosclerosis | 229 (1.1%) |
| Crohn’s disease | 80 (0.4%) |
| Dementia | 348 (1.7%) |
| Depressive disorder | 2543 (12.6%) |
| Diabetes mellitus | 5191 (25.8%) |
| Gastroesophageal reflux disease | 815 (4.0%) |
| Gastrointestinal haemorrhage | 1503 (7.5%) |
| Heart disease | 4872 (24.2%) |
| Heart failure | 755 (3.8%) |
| Hepatitis C | 17 (0.1%) |
| HIV | 6 (0.0%) |
| H pylori gastrointestinal infection | 86 (0.40%) |
| Hyperlipidemia | 2184 (10.9%) |
| Hypertensive disorder | 8551 (42.5%) |
| Ischemic heart disease | 2724 (13.5%) |
| Lesion of liver | 707 (3.5%) |
| Obesity | 722 (3.6%) |
| Osteoarthritis | 5839 (29.0%) |
| Peripheral vascular disease | 501 (2.5%) |
| Pneumonia | 641 (3.2%) |
| Psoriasis | 750 (3.7%) |
| Pulmonary embolism | 498 (2.5%) |
| Renal impairment | 2837 (14.1%) |
| Rheumatoid arthritis | 286 (1.4%) |
| Schizophrenia | 68 (0.3%) |
| Ulcerative colitis | 114 (0.6%) |
| Urinary tract infectious disease | 2945 (14.6%) |
| Venous thrombosis | 1501 (7.5%) |
| Visual system disorder | 8656 (43.0%) |

### **S4: Age standardised by the European Standard Population for incidence rates for CPRD GOLD for pancreatic cancer stratified by sex (red dotted line denotes introduction of Quality and Outcomes Framework (QOF) in 2004)**

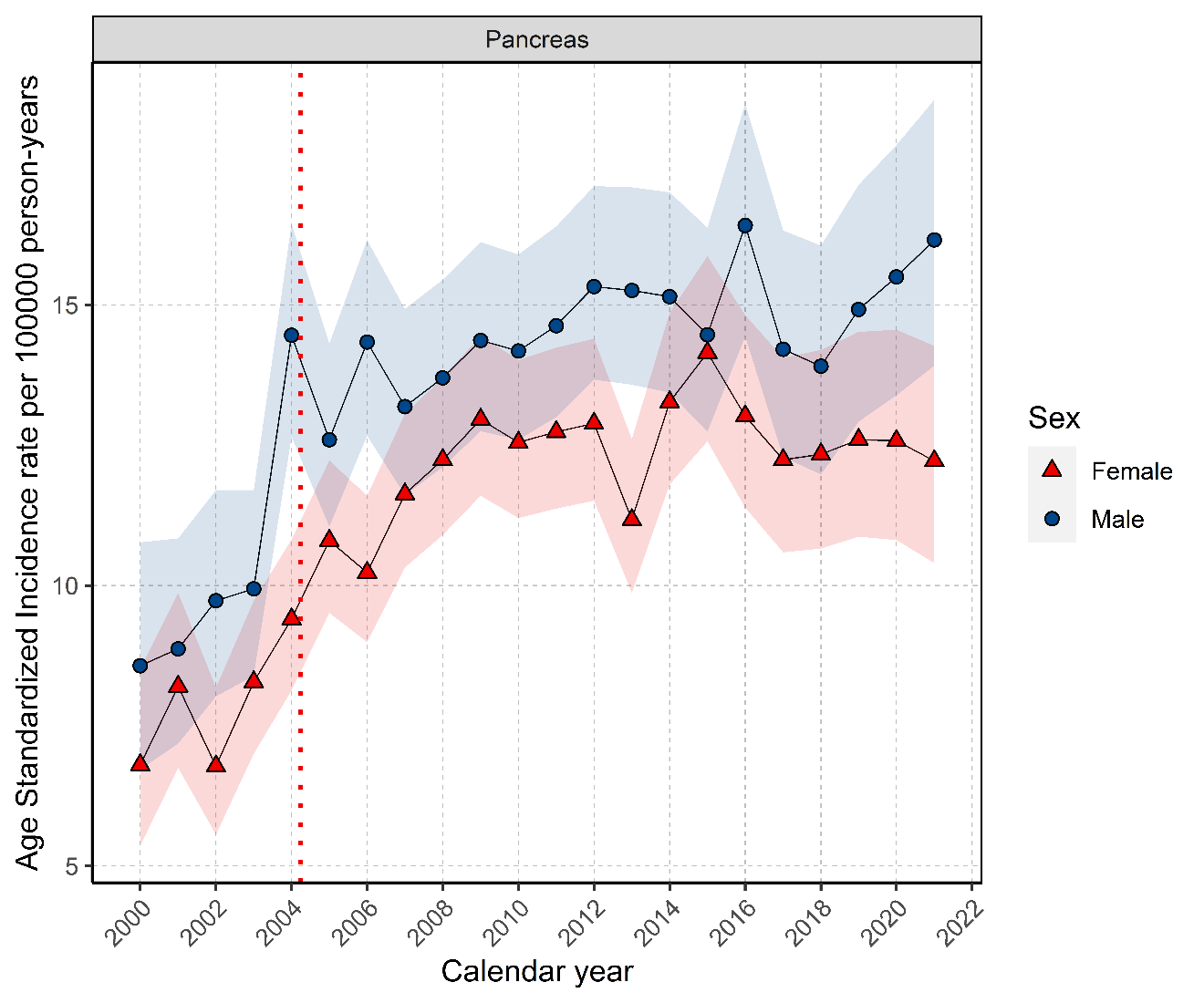

### **S5: Comparison of Age standardised incidence rates for CPRD GOLD versus National Cancer Registration and Analysis Service (NCRAS)**

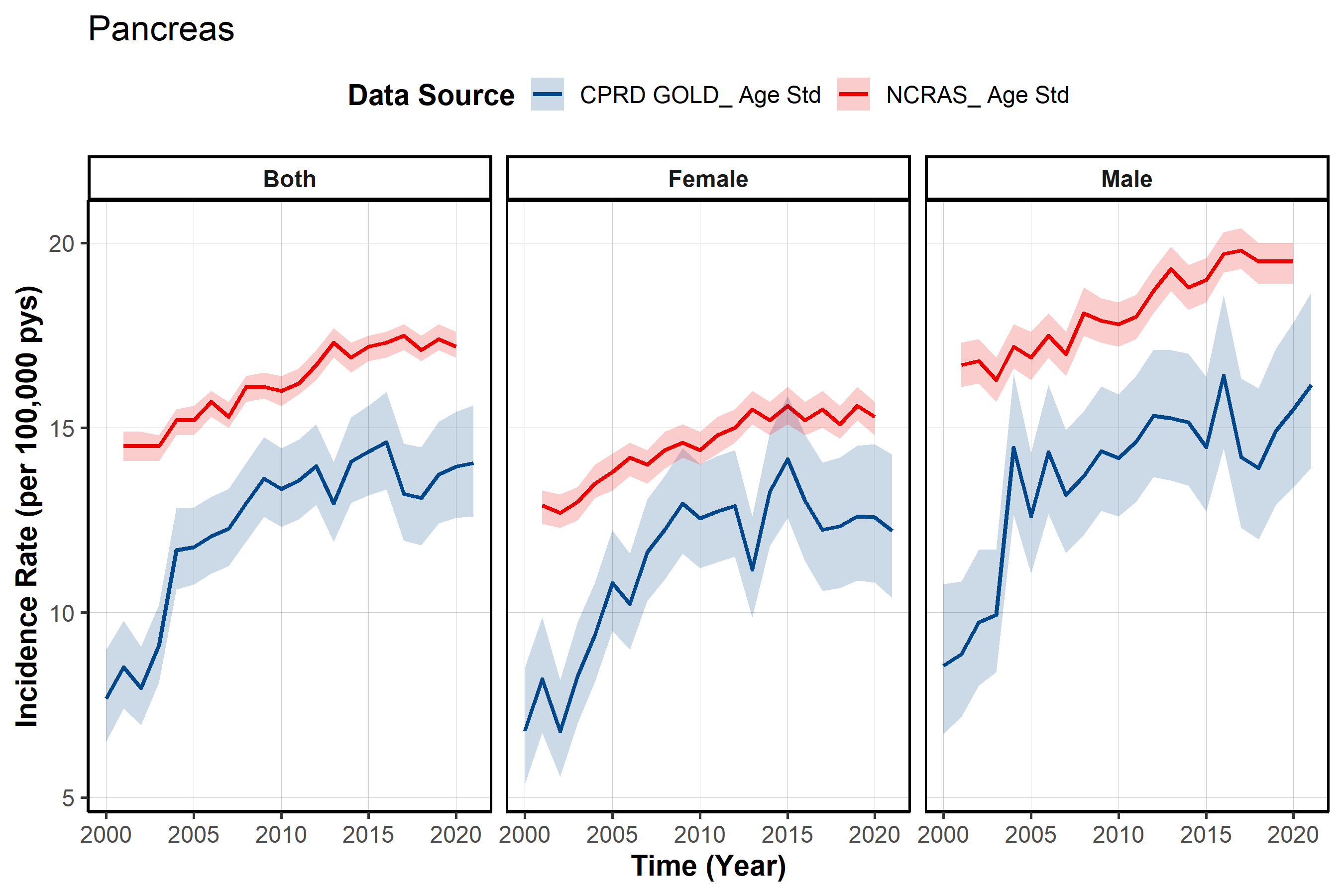

### **S6: Overall incidence rates for pancreatic cancer stratified by database and age group.**

| **Age Group** | **n persons** | **person years** | **n events** | **Incidence (100000 pys)** | **Database** |
| --- | --- | --- | --- | --- | --- |
| 18 to 29 | 9,238,468 | 31,941,580 | 14 | 0.04 (0.02 to 0.07) | Aurum |
| 30 to 39 | 8,292,698 | 32,680,571 | 93 | 0.28 (0.23 to 0.35) |  |
| 40 to 49 | 6,516,206 | 32,926,232 | 656 | 1.99 (1.84 to 2.15) |  |
| 50 to 59 | 5,439,256 | 28,679,169 | 2,214 | 7.72 (7.40 to 8.05) |  |
| 60 to 69 | 4,173,304 | 22,531,700 | 4,873 | 21.63 (21.02 to 22.24) |  |
| 70 to 79 | 3,124,674 | 16,295,746 | 6,661 | 40.88 (39.90 to 41.87) |  |
| 80 to 89 | 1,953,682 | 8,858,773 | 4,748 | 53.60 (52.08 to 55.14) |  |
| 90 + | 667,320 | 2,272,568 | 868 | 38.19 (35.70 to 40.82) |  |
| 18 to 29 | 3,871,136 | 16,018,643 | 6 | 0.04 (0.01 to 0.08) | GOLD |
| 30 to 39 | 3,682,420 | 15,107,587 | 35 | 0.23 (0.16 to 0.32) |  |
| 40 to 49 | 3,247,001 | 16,117,599 | 314 | 1.95 (1.74 to 2.18) |  |
| 50 to 59 | 2,886,092 | 14,726,671 | 1,136 | 7.71 (7.27 to 8.18) |  |
| 60 to 69 | 2,294,832 | 11,907,368 | 2,546 | 21.38 (20.56 to 22.23) |  |
| 70 to 79 | 1,684,767 | 8,428,915 | 3,316 | 39.34 (38.01 to 40.70) |  |
| 80 to 89 | 1,025,886 | 4,433,245 | 2,337 | 52.72 (50.60 to 54.90) |  |
| 90 + | 323,191 | 966,580 | 426 | 44.07 (39.99 to 48.46) |  |

**Pys person years**

### **S7: Annualised incidence rates for pancreatic cancer stratified by database, sex, and age group.**

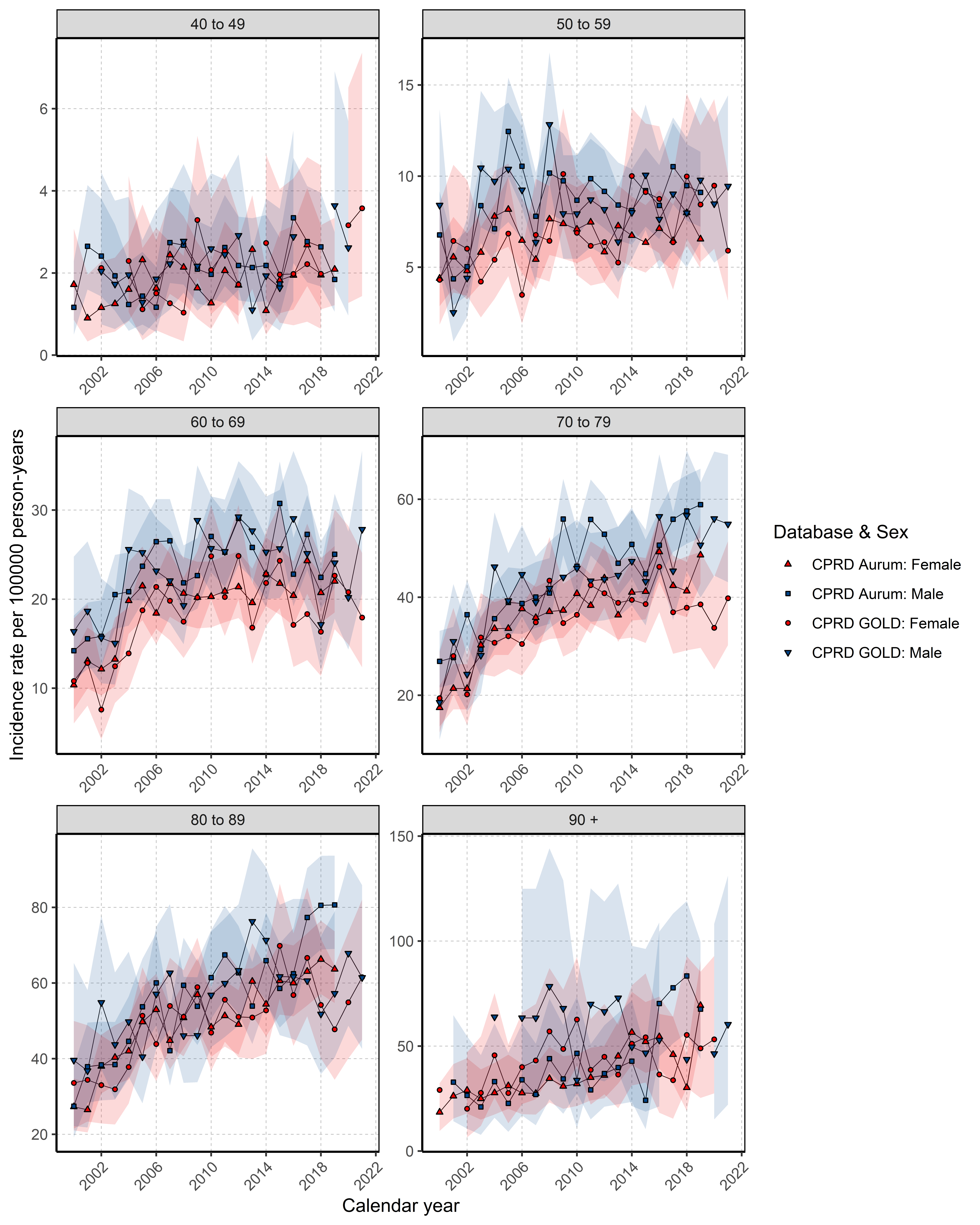

### **S8: Annualised prevalence for pancreatic cancer stratified by database, sex, and age group.**

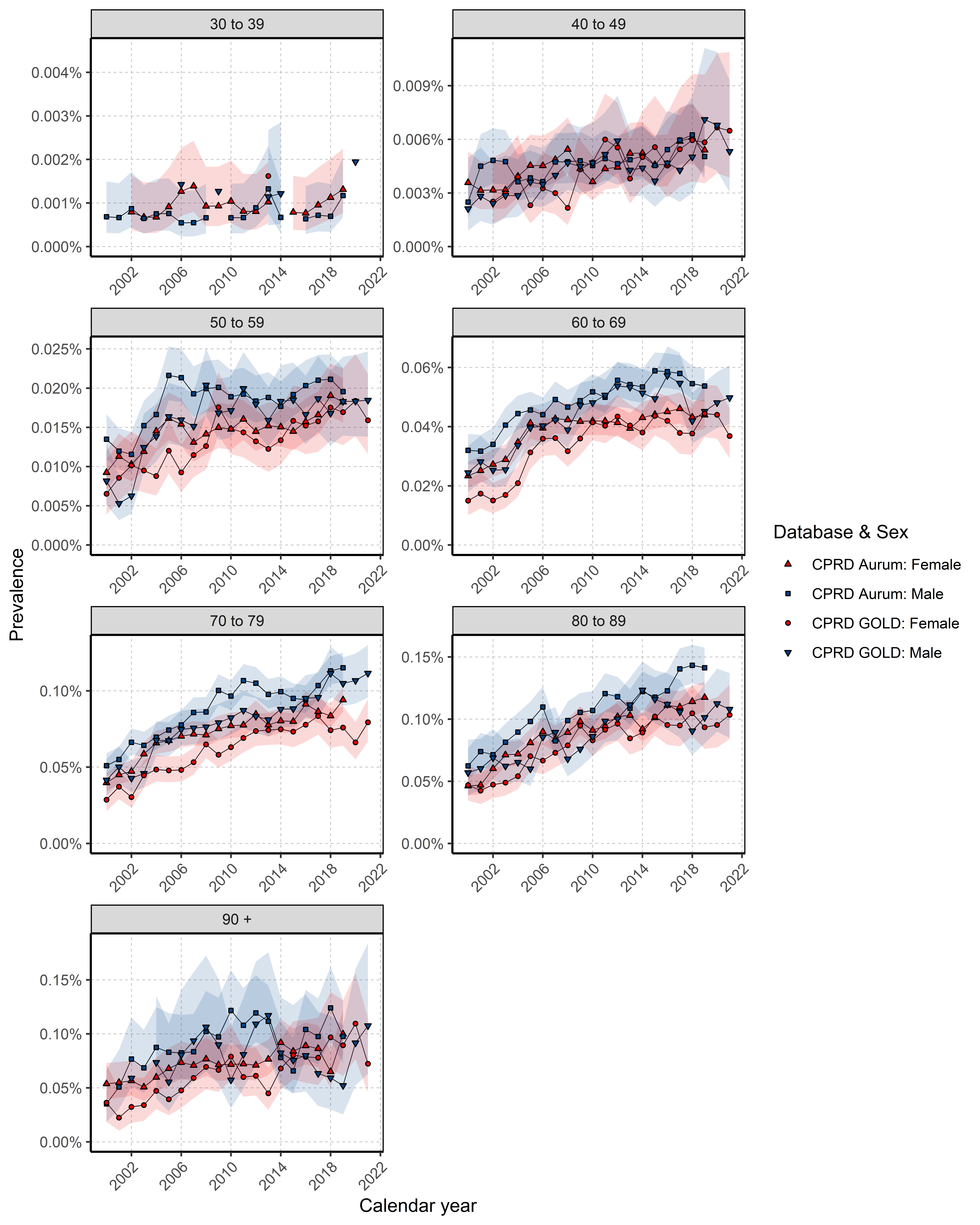

### **S9: Kaplan-Meier survival curve of pancreatic cancer by database and sex**

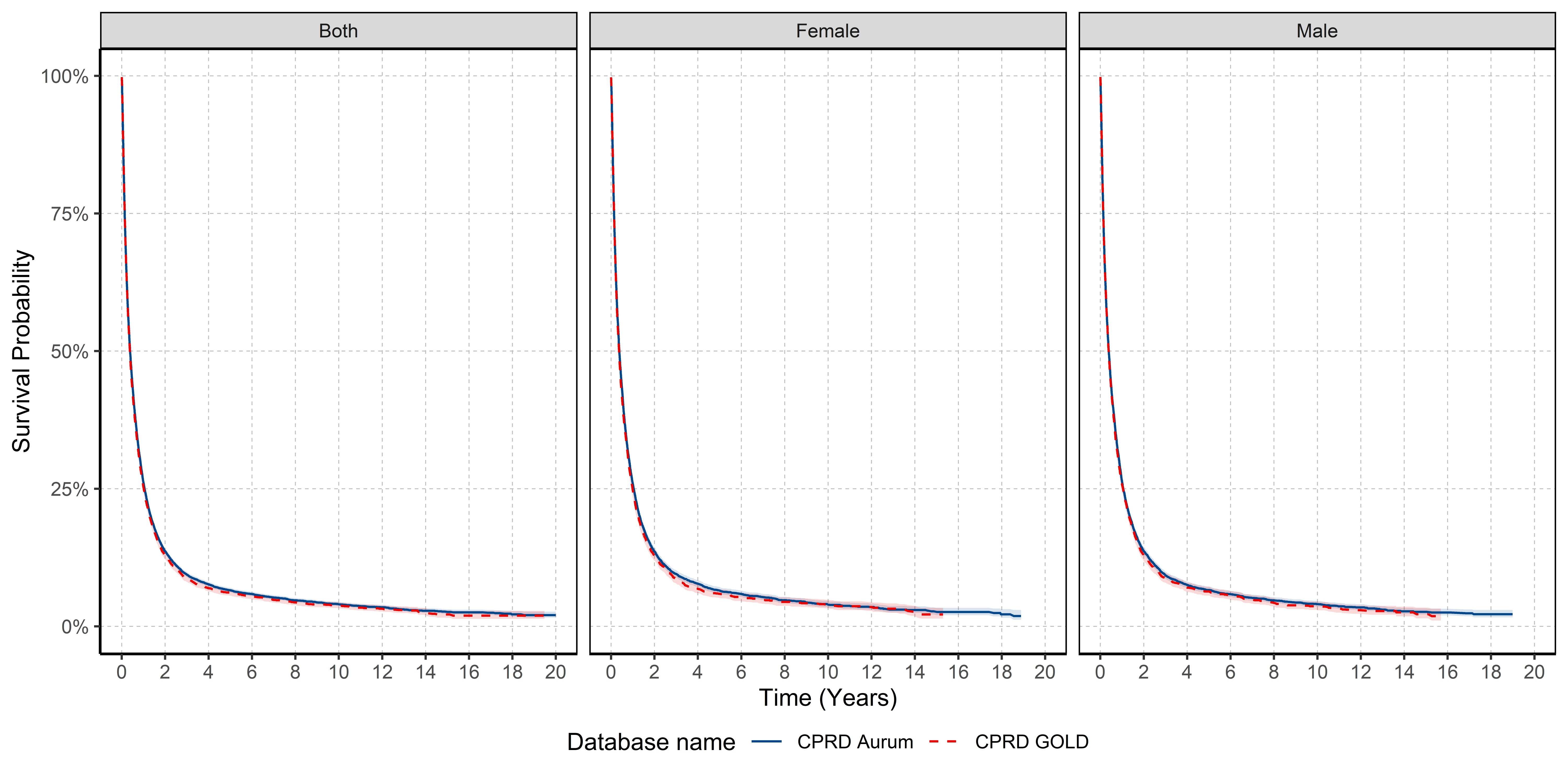

### **S10: Survival (%) after one, five and ten years after pancreatic cancer diagnosis stratified by database and sex.**

| **Time** | **Sex** | **% Survival (95% CI)** | **Database** |
| --- | --- | --- | --- |
| 1 | Male | 26.3 (25.4 - 27.3) | Aurum |
| 5 |  | 6.6 (6.1 - 7.2) |  |
| 10 |  | 4.0 (3.6 - 4.6) |  |
| 1 | Female | 25.9 (25.0 - 26.9) |  |
| 5 |  | 6.5 (6.0 - 7.1) |  |
| 10 |  | 4.0 (3.4 - 4.6) |  |
| 1 | Male | 25.8 (24.5 - 27.1) | GOLD |
| 5 |  | 6.3 (5.6 - 7.2) |  |
| 10 |  | 3.7 (3.0 - 4.5) |  |
| 1 | Female | 24.7 (23.5 - 26.0) |  |
| 5 |  | 5.9 (5.16 - 6.8) |  |
| 10 |  | 4.0 (3.25 - 4.8) |  |

CI: confidence interval

### **S11: Survival (%) after one and five years after pancreatic cancer diagnosis stratified by sex and calendar year**

| **Time (years)** | **% Survival (95% CI)** | **Sex** | **Calendar Year** |
| --- | --- | --- | --- |
| 1 | 22.5 (20.2 - 25.1) | Both | 2000 to 2004 |
|  | 22.9 (19.7 - 26.5) | Male |  |
|  | 22.2 (19.0 - 25.8) | Female |  |
|  | 23.3 (21.7 – 25.0) | Both | 2005 to 2009 |
|  | 23.7 (21.4 - 26.1) | Male |  |
|  | 22.9 (20.7 - 25.3) | Female |  |
|  | 26.0 (24.4 - 27.7) | Both | 2010 to 2014 |
|  | 27.0 (24.7 - 29.4) | Male |  |
|  | 25.1 (22.9 - 27.5) | Female |  |
|  | 28.4 (26.5 - 30.5) | Both | 2015 to 2019 |
|  | 29.0 (26.3 - 32.0) | Male |  |
|  | 27.8 (25.2 - 30.8) | Female |  |
|  | 25.1 (21.4 - 29.3) | Both | 2020 to 2021 |
|  | 23.8 (19.0 - 29.9) | Male |  |
|  | 26.4 (21.2 - 32.8) | Female |  |
| 5 | 6.34 (5.04 - 7.97) | Both | 2000 to 2004 |
|  | 6.12 (4.41 - 8.51) | Male |  |
|  | 6.57 (4.77 - 9.05) | Female |  |
|  | 5.46 (4.62 - 6.46) | Both | 2005 to 2009 |
|  | 5.33 (4.20 - 6.78) | Male |  |
|  | 5.64 (4.47 - 7.12) | Female |  |
|  | 6.01 (5.08 - 7.12) | Both | 2010 to 2014 |
|  | 6.63 (5.32 - 8.27) | Male |  |
|  | 5.34 (4.11 - 6.95) | Female |  |
|  | 6.79 (5.54 - 8.32) | Both | 2015 to 2019 |
|  | 7.71 (5.91 - 10.1) | Male |  |
|  | 5.92 (4.34 - 8.10) | Female |  |
